# Longitudinal Association of Mid-Life Ten Year Cardiovascular Disease Risk Score with Brain Biomarkers of Alzheimer’s Disease, Neurodegeneration and White Matter Hyper Intensities in Cognitively Unimpaired Older Adults: Heart SCORE Brain Study

**DOI:** 10.1101/2024.01.24.24301752

**Authors:** Anum Saeed, Yue Fang Chang, Justin Swanson, Michael Vu, Mark Mapstone, Victor L Villemagne, Beth E. Snitz, Sarah K Royse, Brian Lopresti, Howard J. Aizenstein, Minjie Wu, Kevin Kip, Steven E. Reis, Oscar Lopez, Ann Cohen

## Abstract

**Introduction:** Atherosclerotic cardiovascular disease (ASCVD) risk factors in mid-life have been associated with cognitive decline and late-life dementia. However, the role of these risk factors in preclinical Alzheimer’s disease (AD) pathophysiology remains elusive. We investigated whether mid-life 10-year pooled cohort equations (PCE) based ASCVD risk is associated with late-life amyloid, tau, neurodegeneration [AT(N)] measures and white matter hyperintensities (WMHI).

**Methods:** Participants enrolled in the Heart Strategies Concentrating on Risk Evaluation (Heart SCORE) study between 2003-2005 (mid-life) and underwent brain MRI and PET scans in 2018-2022 (age >65 years, late-life) to detect and quantify amyloid (A, PiB-PET) and tau (T, Flortaucipir (FTP) PET) deposition, cortical thickness (N) and white matter hyperintensities (WMHIs). Mid-life PCE ASCVD risk was categorized as*; borderline (5%-7.4%), intermediate (7.5%-<15%), or high (≥15%*). Association of midlife ASCVD risk HR (5% CI) was assessed using logistic and linear regressions with A, T, or N and chi square beta coefficients for WMHI in latelife.

**Results:** Over a ∼16y follow up, in 135 participants (mean age 73y), A and T showed no significant association with mid-life ASCVD risk. Neurodegeneration had a graded association with mid-life ASCVD risk categories (*OR_ASCVD high vs low risk%_ 6.98 [2.44-19.95]; p<0.05*) driven by self-identified Black race and age. In a subset n=60, ASCVD risk score was also associated with WMHIs ((β=*0.42 ± 0.22; p=0.05)* in a model adjusted for inflammation and education.

**Conclusions:** In this asymptomatic, diverse cohort, 10y ASCVD risk was predictive of late-life neurodegeneration and white matter hyperintensities but not amyloid or tau. These data suggest that ASCVD risk factors in midlife may lead to a state of vulnerability (through increased neurodegeneration and white matter hyperintensities) which may progress to cognitive decline and dementia. Further mechanistic studies are warranted to test this hypothesis.

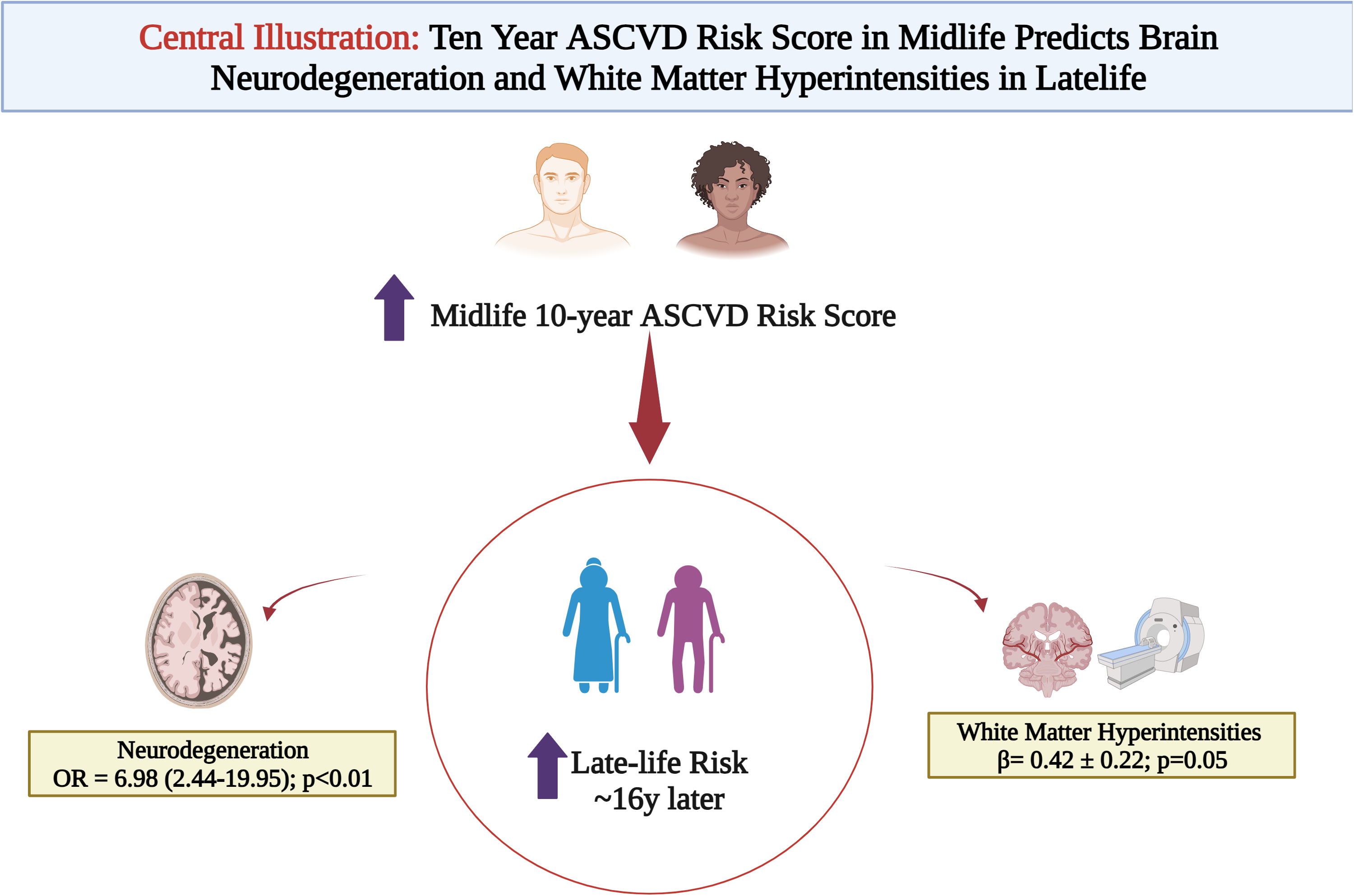

## Introduction

Alzheimer’s disease (AD) and AD-Related Dementias (ADRD) are a major public health challenge in the United States with projected increases to 9.3 million by 2060 (1). Development of disease-modifying therapies are challenging due, in part, by the long preclinical window of AD. The preclinical window of AD can begin in mid-life, providing a critical window for identification and optimization of AD risk factors.

In the National Institute on Aging and Alzheimer’s Association Research Framework, AD is defined by its underlying pathologic processes that can be documented by postmortem examination or *in vivo* by biomarkers. The diagnosis is not based on the clinical consequences of the disease (i.e., symptoms/signs) in this research framework, which shifts the definition of AD in living people from a syndromal to a biological construct. The research framework focuses on the diagnosis of AD with biomarkers in living persons. Biomarkers are grouped into those of β amyloid (A) and tau (T) deposition, and neurodegeneration (N) or collectively identified as AT(N). This classification system groups different biomarkers (imaging and biofluids) by the pathologic process underlying each measures.

Neurodegenerative changes (N) in the absence of amyloid and tau, in addition to white matter hyperintensities (WMHIs) are suggested to be linked with various forms of cognitive decline though their involvement in AD onset remains elusive. Both WMHIs and neurodegenerative changes, like ASCVD can present years before cognitive decline (2, 3). Hypertension, inflammation and dyslipidemias, risk factors for ASCVD, have been linked with neurodegeneration and WMHIs in individuals with cerebrovascular pathology and without (4, 5) (6) (7).

In the United States, 10-year ASCVD risk assessed calculated by the pooled cohort equations (PCE) is used as a guiding tool to identify individuals are risk for ASCVD events (8). In the 2013 and 2018 American Heart Association and American College of Cardiology’s cholesterol guidelines, the PCE derived 10-year risk was recommended to guide the use of statin therapy in primary prevention. Although the PCE predicts ASCVD events, its risk prediction for dementia or AD pathology has not yet been assessed. This study investigated the association of PCE based ASCVD risk score with cerebrovascular white matter damage as WMHIs, neurodegeneration and brain biomarkers of AD in a biracial cohort of community dwellers with preserved cognition over a ∼16 year follow up period.

## Methods

The Heart Strategies Concentrating on Risk Evaluation (Heart SCORE) study originally enrolled 1,949 participants aged 45-75 years at baseline entry between 2003-2005 (∼44% racialized as Black, ∼66% women) to examine racial disparities in the prevalence and outcome of ASCVD (NCT00143923) (9, 10). The cohort underwent a baseline evaluation of demographic and psychosocial characteristics and serologic markers including all the variables of the PCE derived ASCVD risk (i.e age, sex, self-reported race, total cholesterol, high density lipoprotein cholesterol (HDL-c), triglycerides, diabetes, hypertension medications, systolic blood pressure and smoking history). From the original Heart SCORE study, a subset of participants who are currently undergoing late-life neuroimaging and neurocognitive testing, were recruited for this analysis. All participants underwent a baseline Montreal Cognitive Assessment (MoCA) score >20; those who met criteria for dementia at a Consensus Diagnostic Conference were excluded from the current study.

### Brain MRI scans

The MR scanning was completed on a Siemens Prisma 3-T scanner, equipped with 64-channel systems with Connectome level gradients operating at 80mT/m (11). Briefly, the following sequences were obtained from all participants: MP-RAGE (Magnetization Prepared - Rapid Gradient Echo), axial gradient recalled echo T2-weighted imaging, axial T2 fluid-attenuated inversion recovery (FLAIR).

We processed T1 images with FreeSurfer version 7.1 to define regions-of-interest (ROIs) and total intracranial volume (ICV)(12). We applied the Imperial College London Clinical Imaging Center (CIC) atlas to better delineate functional subdivisions of the striatum(13). All parcellations were visually inspected prior to analysis. We calculated a composite measure of medial temporal lobe (MTL) cortical thickness (mm) as the surface area-weighted average of FreeSurfer ROI-derived cortical thickness in the fusiform gyrus, entorhinal cortex, inferior and middle temporal gyri (15). Participants were categorized as N+ if their MTL cortical thickness </= 2.8 OR 2.7 (14). WMHI volume was derived from T2 FLAIR images using a quantitative computer-aided segmentation program to measure the volumetric burden (cm3) of WMHI volume (15).

Prior to the PET imaging session, a T1-weighted structural MR series (MPRAGE) was acquired using a 3T Siemens PRISMA scanner with a 1 mm isotropic voxel size. Each participant’s MR image was parcellated into a set of regions of interest (ROIs) using the default FreeSurfer v5.3 ((12)) pipeline and Desikan-Killiany atlas with the exception of striatal subregions, which were substituted by components from the Imperial College London Clinical Imaging Centre (CIC) atlas (Tziortzi et al. 2011) as previously described (16). All FreeSurfer ROIs were visually inspected and manually edited where appropriate.

An AD-signature composite cortical thickness index was derived based on a surface-area weighted average of the mean cortical thickness of four FreeSurfer ROIs: entorhinal, inferior temporal, middle temporal, and fusiform. Abnormal composite cortical thickness indicating neurodegeneration (N+) in the ATN framework was defined as < 2.7 mm based on a sensitivity threshold (14, 17).

### PET Imaging with Pittsburgh Compound B (PiB)

Detailed methods for PiB PET have been published (18–20). Brain Aβ burden was measured via [11C]Pittsburgh Compound-B (PiB) (21) on either a Siemens Biograph mCT PET/CT scanner (Siemens Healthcare GmbH, Erlagen, Germany) or a Siemens ECAT Exact HR+ scanner (Siemens Healthcare GmbH, Erlagen, Germany). We measured brain tau load using [18F]Flortaucipir (FTP) on the same Siemens Biograph mCT PET/CT described above. Briefly, at separate PET visits, participants received slow bolus injections of either PiB (∼15 mCi) or FTP (∼10 mCi). After a tracer-appropriate delay (PiB: 25-minutes; FTP: 75-minutes), participants were positioned in the scanner for a transmission scan for attenuation correction. PiB PET studies were then acquired 50-70 minutes post-injection; FTP PET studies were acquired 75-105 minutes post-injection.

Each PiB and FTP scan was visually examined and, if necessary, corrected for interframe motion prior to analysis. We created a single PET frame by averaging the images over tracer-specific post-injection intervals (PiB: 50-70 minutes; FTP: 80-100 minutes). Using PMOD version 4.1 (http://www.pmod.com), we co-registered each single-frame PET image to its corresponding MRI in native space and sampled the images with FreeSurfer and CIC atlas ROIs.

Global Aβ burden for each participant was measured as a single, global standardized uptake value ratio (SUVR), calculated as the volume-weighted average of PiB radioactivity in nine FreeSurfer-defined ROIs, normalized to cerebellar gray matter (22).We considered participants to be A+ if their global PiB SUVR >/= 1.3 (22).

We used meta-temporal SUVR as a measure of tau burden by calculating the volume-weighted average of FTP radioactivity in FreeSurfer-defined entorhinal cortex, hippocampal gyrus, fusiform, inferior temporal gyrus, and middle temporal gyrus, normalized to cerebellar gray matter (22). Participants were considered T+ if their meta-temporal FTP SUVR >/= 1.18.

### Covariates and PCE 10-year ASCVD Risk

Baseline characteristics of the study population were defined using data at the entry into the Heart SCORE cohort. These included sociodemographic data (age, sex, and race/ethnicity, low-density lipoprotein cholesterol, and the remaining pooled cohorts equation (PCE) variables including; total cholesterol, high-density lipoprotein cholesterol, treatment for hypertension, systolic blood pressure, current smoking status, and diabetes. The cohort was divided into categories of PCE-derived 10-year risk (8, 23) that is, *low-risk (<5%); borderline risk (5%–7.4%); intermediate risk (7.5%–<15%), and high risk (≥15%)*.

Detailed methods have been described elsewhere (9). Briefly, lipid measurements were performed on fasting plasma samples that were stored at −70°C with ethylenediaminetetraacetic acid as the anticoagulant. Plasma total cholesterol, high-density lipoprotein cholesterol, and triglycerides were measured using enzymatic measures (24). Low-density lipoprotein cholesterol was calculated using the Friedewald equation (25). Diabetes mellitus was defined as a fasting glucose level ≥126 mg/dL, a self-reported physician diagnosis of diabetes, or use of diabetes medications. Hypertension in the Heart SCORE study is defined as systolic blood pressure ≥140 mm Hg or diastolic blood pressure ≥90 mm Hg or use of blood pressure–lowering therapies. Consistent with the PCE, smoking history was categorized as current or not current (a composite of never and former smoking). Inflammatory burden at baseline was assessed by measuring high sensitivity C reactive protein (hsCRP) on participants’ plasma samples.

### Arterial Stiffness

Arterial stiffness and wave reflection were measured at Heart SCORE study entry (mid-life) using the Endo-PAT device and expressed as heart rate-adjusted augmentation index (AI75). (26) Higher AI75 denoted higher arterial stiffness and wave reflection.

## Statistical Analysis

Baseline descriptive statistics of the study sample, such as the minimum, maximum, median, and mean for each continuous variable and frequency table for each categorical variable, were initially analyzed to summarize the data and detect outliers and missing values. Missing data were uncommon. Descriptive characteristics are presented as mean and SD for continuous variables, and frequencies and proportions for categorical variables. The difference of mean across the ASCVD groups is assessed by 1-way ANOVA, while the difference of frequencies is compared using the χ^2^ test.

White matter hyperintensities volume was natural log-transformed to adjust for skewness. We used linear and logistic regression models to estimate the association of the ten year ASCVD risk score with WMHI volume, brain amyloid deposition and neurodegeneration. We evaluated midlife 10 year ASCVD risk score, as continuous and categorical variables. All models were fully adjusted for education level and hsCRP measures. Further, WMHI were adjusted for total intracranial volume.

In an exploratory analysis, we also assessed association of arterial stiffness (AI75) in midlife as a continuous variable with latelife WMHIs.

## Results

### Baseline Characteristics

#### Participants with PET imaging

A subset of 145 Heart SCORE participants with PET imaging were included in the current analysis out of whom 68.3% were females and ∼30% of self-identified Black race (**Table 1a**). Overall, 35.2% of the participants had baseline hypertension and ∼38% were categorized as high risk according to the 10-year ASCVD risk in midlife. In race-stratified baseline characteristic, more Black participants had baseline hypertension (54.8%) and diabetes (7%). More Black participants (∼62%) were noted to have a higher 10-year ASCVD risk (>7.5%) in midlife compared to White (31%) counterparts (**Table 1a**).

**Table 1A:**
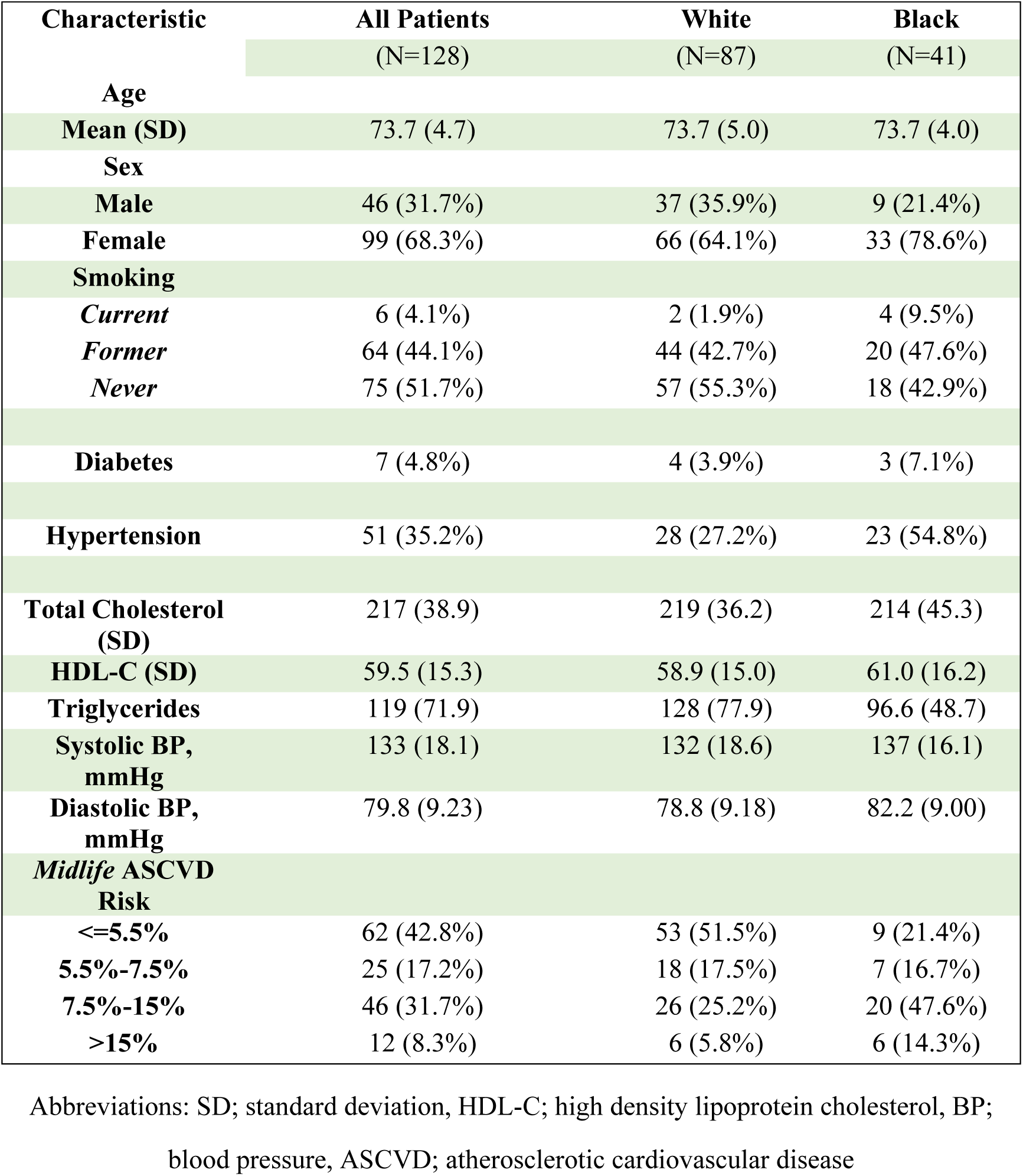
Baseline Characteristics of the Heart SCORE participants undergoing Brain PET Imaging.

#### Participants with White Matter Hyperintensities

As subset of 60 participants included from the Heart SCORE brain MRI study were included in the current analysis (**Table 1b**). Mean age at baseline was 59.7 ± 4.4 years and ∼60% were females. Like the overall cohort, 30% of participants who underwent brain MRI self-identified as Black and ∼38% had hypertension at baseline. The mean ASCVD risk score for this subset was 8±0.06% (**Table 1b**).

**Table 1B:** Baseline Characteristics of the Heart SCORE participants undergoing Brain MRI Imaging.

| Variable | Heart SCORE Study Cohort |
| --- | --- |
| **Total Population** | 60 |
| **Age, yrs**<br>**(mean±SD)** | 73.1 ±4.6 |
| **Female**<br>**n (%)** | 36 (60%) |
| **White**<br>**n (%)** | 41 (68.3%) |
| **Black**<br>**n (%)** | 19(31.7%) |
| **>College Education**<br>**n (%)** | 37 (38%) |
| **Hypertension**<br>**n (%)** | 23 (38%) |
| **Smoking**<br>**n (%)** | 3 (5%) |
| **Diabetes Mellitus**<br>**n (%)** | 2 (3.3%) |
| **Total Cholesterol mg/dL**<br>**(mean±SD)** | 218.5±41.1 |
| **HDL-C mg/dL**<br>**(mean±SD)** | 58.2±15.5 |
| **Systolic BP mmHg**<br>**(mean±SD)** | 134.2±19.1 |
| **hsCRP** | 1.5 (0.7, 4.2) |
| **ASCVD risk score** | 0.08±0.06 |

Abbreviations: SD; standard deviation, HDL-C; high density lipoprotein cholesterol, BP; blood pressure, ASCVD; atherosclerotic cardiovascular disease

#### Association of Midlife ASCVD Risk and Latelife Brain Biomarkers of Neurodegeneration using cortical thickness

Neurodegenerative (N) biomarker had a graded association with increasing mid-life ASCVD risk categories *(OR_ASCVD high vs low risk%_ 6.98 [2.44-19.95]; p<0.05*) while no significant association was found with PiB uptake (A) on PET **(Table 2)** after a mean 16y follow up. In fully adjusted models, the association of neurodegeneration brain biomarker with the 10 year ASCVD risk was mainly driven by Black race (*RR 3.18 [1.77-5.12]*), age (*RR 1.84 [1.35-2.42]*) and aspirin use *(RR 2.10 [1.34-3.26]; all p<0.05* **(Supplemental Table II).** Baseline education level and hypertension were not associated with late-life neurodegeneration.

Given the significantly higher risk of neurodegeneration biomarker in Black participants, we conducted a race-stratified regression analysis (**Table 2a**). A graded risk was observed across the ASCVD risk score amongst Black participants as well as White. However given a limited number of Black participants, we did not observe significant associations. Amongst White participants, midlife 10-year risk of >15% had the highest risk of neurodegeneration *(RR 8.46 [1.89-37.88];p=0.01)* (**Table 2a**).

**Table 2a:** Midlife 10 year ASCVD Risk and Association with Brain Biomarkers of Alzheimer’s Disease on PET Imaging.

| ASCVD Risk Score | Brain Amyloid Deposition<br>OR (95% CI) | Neurodegeneration<br>OR (95% CI) |
| --- | --- | --- |
| ≤5.5% | Reference | Reference |
| 5.5% - <7.5% | 0.65 (0.17-2.08) | 2.73 (0.96-7.79)~ |
| 7.5% - <15% | 0.42 (0.13-1.20) | **3.27 (1.33-8.03)\*** |
| ≥ 15% | 1.14 (0.23-4.46) | **6.98 (2.44-19.95)\*** |
| ~ p-value < .1; * p-value < .05 |  |  |

Abbreviations: ASCVD; atherosclerotic cardiovascular disease, RR; relative risk, CI; confidence interval.
Amyloid deposition by Pittsburgh compound B uptake on imaging.

#### Association of Midlife ASCVD Risk and Latelife Brain Biomarkers of AD

We did not find a statistically significant association of midlife 10-year ASCVD risk with brain biomarkers of AD (A or T). Although there was a trend noted towards higher risk of amyloid (A) deposition (OR 1.14 [0.23-4.46)], it did not reach statistical significance.

#### Association of Midlife ASCVD Risk and Latelife White Matter Hyperintensities on Brain MRI Imaging

In the continuous linear regression, latelife WMHI lesion burden significantly correlated with midlife ASCVD 10-year risk (β=*0.51 ± 0.21; p=0.02).* When these data were adjusted for hsCRP and education level <college, this association was attenuated (*0.42 ± 0.22, p=0.05*) (**Table 3a**). There was a significant negative association with WMHI burden in latelife with midlife elevated hsCRP *(β= -0.25 ± 0.11; p=0.03)* while no significance was observed with education level. We observed a graded association with risk of WMHI burden in participants with moderate to high midlife 10 year ASCVD risk when stratified by categories of risk (**Table 3b**). In model 2 (adjusted for hsCRP and education level), the association WMHIs with midlife ASCVD risk of >15% was no longer statistically significant; likely driven by inflammation burden (*hsCRP β= -0.28 ± 0.11; p=0.02).* In the exploratory analysis, there was a signal of association between midlife vascular stiffness and late-life WMHIs in this cohort (*β= 0.016 ± 0.009; p=0.06)* largely driven by presence of hypertension (*β=* 0.88 *±* 0.39*; p=0.03)*.

**Table 2b:** Midlife 10 year ASCVD Risk and its Association with Neurodegeneration Stratified by Race.

| ASCVD Risk Score | White |  | Black |  |
| --- | --- | --- | --- | --- |
|  | RR (95% CI) | P value | RR (95% CI) | P value |
| ≤5.5% | Reference |  | Reference |  |
| 5.5% - <7.5% | 2.32 (0.52-10.35) | 0.27 | 2.03 (0.45-9.06) | 0.36 |
| 7.5% - <15% | 3.32 (0.94-11.77) | 0.06 | 1.46 (0.40-5.40) | 0.57 |
| ≥ 15% | **8.46 (1.89-37.88)** | **0.01** | 2.55 (0.57-11.42) | 0.22 |

Abbreviations: ASCVD; atherosclerotic cardiovascular disease, RR; relative risk, CI; confidence interval

**Table 3:** Association of 10 Year Atherosclerotic Cardiovascular Risk Score with White Matter Hyper intensities on Brain MRI.

a) **Linear Regression with ASCVD as Continuous Variable**
| ASCVD Risk Score | Estimate | SE | p-value |
| --- | --- | --- | --- |
| **Model 1** | **0.51** | **0.21** | **0.02** |
| **Model 2** | **0.42** | **0.22** | **0.05** |

\* Regression coefficient was for 1SD (=0.089) increment.
\* Model 1= uncorrected
\*Model 2= adjusted for education and hsCRP

b) **Linear Regression with ASCVD Score as Categorical Variable**
| Model | ASCVD risk score Categories | Estimate | SE | p-value |
| --- | --- | --- | --- | --- |
| **Model 1** | **5.5%-<7.5%** | 0.25 | 0.50 | 0.62 |
|  | **7.5% - <15%** | 0.66 | 0.35 | 0.06 |
|  | **≥ 15%** | **0.98** | **0.50** | **0.05** |
| *<br>**Model 2** | **5.5%-<7.5%** | 0.23 | 0.51 | 0.65 |
|  | **7.5% to <15%** | **0.73** | **0.34** | **0.04** |
|  | **>= 15%** | 0.67 | 0.52 | 0.21 |

Model 1= uncorrected
Model 2= adjusted for education and hsCRP

**Table 4:**
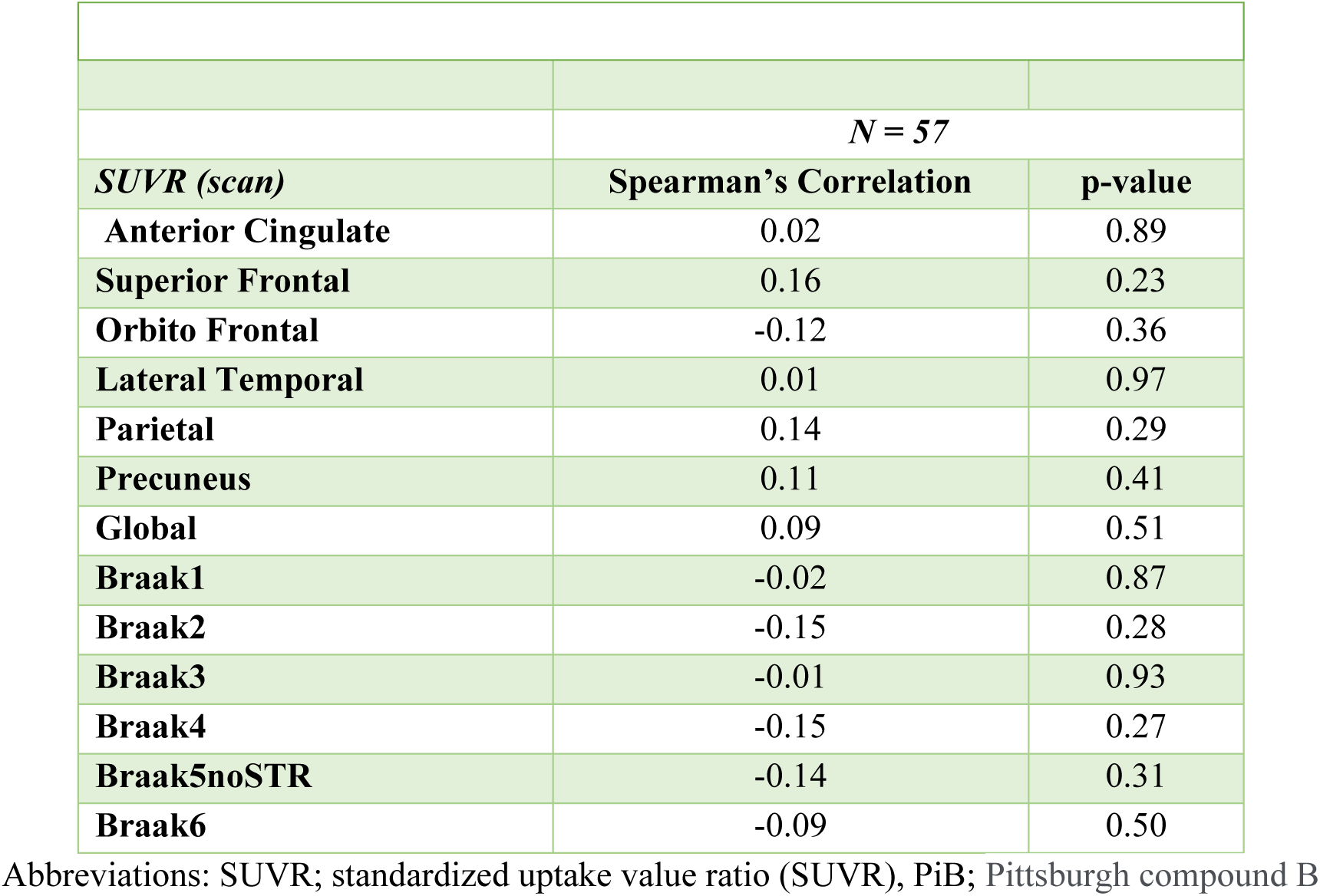
Spearman Correlation of PiB outcomes and normalized white matter hyperintensities volume.

|  | *N = 57* |  |
| --- | --- | --- |
| *SUVR (scan)* | **Spearman's Correlation** | **p-value** |
| **Anterior Cingulate** | 0.02 | 0.89 |
| **Superior Frontal** | 0.16 | 0.23 |
| **Orbito Frontal** | -0.12 | 0.36 |
| **Lateral Temporal** | 0.01 | 0.97 |
| **Parietal** | 0.14 | 0.29 |
| **Precuneus** | 0.11 | 0.41 |
| **Global** | 0.09 | 0.51 |
| **Braak1** | -0.02 | 0.87 |
| **Braak2** | -0.15 | 0.28 |
| **Braak3** | -0.01 | 0.93 |
| **Braak4** | -0.15 | 0.27 |
| **Braak5noSTR** | -0.14 | 0.31 |
| **Braak6** | -0.09 | 0.50 |

Abbreviations: SUVR; standardized uptake value ratio (SUVR), PiB; Pittsburgh compound B

## Discussion

In a longitudinal cohort of community dwellers without cognitive impairment we present three key findings; 1) midlife10-year ASCVD risk was significantly associated with white matter hyperintensities and neurodegeneration, while not with amyloid and tau, on brain imaging obtained more than 15 years later, 2) this association was graded across increasing categories of risk score, and; 3) association of ASCVD risk and neurodegeneration was primarily driven by presence of self-reported Black race. To our knowledge, this is the first study evaluating midlife ASCVD risk score and its long-term association with WMHIs, neurodegeneration and Alzheimer’s disease (AD) imaging biomarker burden in the same cohort of cognitively preserved persons.

Our findings are meaningful in several ways. The association of a routinely implemented 10y ASCVD risk score may be an important risk stratification tool for early intervention in prevention of WMHIs and neurodegeneration – both of which may induce a state of “brain vulnerability” and progression to dementia and AD (27). The ASCVD risk score is an established predictor of ASCVD events within the Heart SCORE population (10, 28, 29) and other cohorts across the United States (30, 31). This score is routinely implemented in clinical practice by clinicians (such as primary care physicians, geriatricians, internists and cardiologists) to identify high risk individuals who will benefit from aggressive preventive interventions including statin therapy initiation. (32) Thus, to utilize a common tool for risk stratification for future ASCVD and a “*vulnerable*” brain state is an important advance in risk stratification and patient-physician discussion on preventive care. Havenon et al., recently reported the association of ASCVD risk with elevated WMHI burden and progression in their preliminary work. (33) Individually, hypertension and other cardiac risk factors have been associated with WMHI and neurodegeneration in several cross-sectional analysis (34) (35). However, longitudinal prediction of WMHIs, neurodegeneration and brain AD biomarkers with a routinely used clinical tool has not been performed in a cognitively persevered and diverse community cohort.

The significance of WMHIs as an important neuroimaging characteristic in has been established recently in those with autosomal dominant AD (ADAD). (36). In individuals with ADAD and cognitive impairment, Schoemaker et al., found that WMHI volume was higher compared to both cognitively unimpaired ADAD and non-carriers. Conversely, cortical amyloid burden on PET and cerebrospinal fluid (CSF) phosphorylated tau levels were elevated, and CSF amyloid levels were reduced in both cognitively unimpaired and impaired mutation carriers, relative to non-carriers. Rizvi et al., have also shown that WMHIs were associated with cortical thinning in the right frontal/parietal regions over a span of 4 years. (37) These patterns of atrophy align with the neurodegenerative changes commonly observed in AD including the characteristic cortical signature involving the entorhinal cortex and multiple cortical regions.

Cortes-Canteli and coworkers recently showed an association of brain hypometabolism with modifiable cardiovascular risk factors stratified by the Framingham risk score. (38) Indeed risk factors of CVD including hypertension (39), hyperlipidemia (40), metabolic syndrome (41) and cigarette smoking (42) have a very strong association with dementia incidence (43, 44). More recent evidence also links these risk factors for promotion of amyloid and progression of AD pathology. (45–47) Although we did not observe an association of the 10y risk score with brain amyloid and tau in the current analysis; it is plausible that long-term presence of ASCVD risk factors creates a “vulnerable” region for amyloid and tau deposition in the later years and subsequent progression to AD dementia (48–51).

The 10-year atherosclerotic CVD risk score (52), which takes into account presence of modifiable and non-modifiable clinical risk factors has an excellent predictability for ASCVD events. However, during transition from mid-to late-life, the clinical risk factors become more prevalent and are less predictive of clinical disease; thus they likely under-quantify event “risk”. We have previously shown that in older adults, addition of biomarkers of inflammation, myocardial stretch and injury improved ASCVD event prediction (ΔAUC 0.103; continuous NRI 0.484) when added to the 10y risk score clinical variables. (53) Thus, in the current analysis, it is also possible that the 10-year ASCVD risk score is less predictive of AD pathology on PET due to the older age of these individuals. It may be imperative to explore whether ASCVD risk score is associated with elevated AD biomarkers, which can predate the tau and Aβ deposition in PET scans (54–57), thereby more succinctly identifying those at the highest risk of AD.

Our results also signal arterial stiffness and hypertension in midlife as likely primary drivers of WMHIs in midlife as shown previously (58–61). Whereas, neurodegeneration was mainly seen in association with self-reported Black race. Thus, the likely mechanisms that lead both these pathological states in the brain may be different and race-related differences may be at play. Furthermore, since race is a social construct, the impact of disparities in social determinants of health and in lived experience, including inequities in access to medical care need to be assessed as the driver for not only AD but also ASCVD risk.

### Limitations

Our study has some notable limitations. First, Heart SCORE is a highly educated cohort homogeneity in terms of years of education may limit comparisons. However, we expect that high education level would not artificially increase neurodegeneration or WMHIs burden. Second, more precise contributions of genetic influences, lifestyle, stress, and socioeconomic environmental exposure may influence this analysis and we did not test for such interactions. In our cohort, we also observed a low proportion of A and T positive participants thus limiting the assessment of AD pathology. Finally, we measured the 10 year ASCVD risk score at midlife one time. Although the score may be influenced by several cardiovascular risk factors, age is the primary driver of this score and thus overtime it will be increase rather than reverse in a vast majority of individuals. While our study included a good proportion of Black and White individuals, the generalizability to other ethnic minorities and geographic regions may be limited due to lack of representation.

## Conclusion

A key question is if the confluence of ASCVD risk factors, as measured by the 10y risk score, can also quantify future risk of the vulnerable brain by virtue of WMHIs and neurodegeneration? Our current data would point to more exploration of this question in future studies. Statin therapies and other primary preventive measures have shown risk reduction in not only ASCVD events but also dementia incidence (62, 63). However, a comprehensive impact of primordial and primary prevention of ASCVD risk factors on WMHIs, neurodegeneration, AD pathology and overall, cognitive decline remains to be tested in randomized and controlled studies. Further, since this study did not explore the relationship between ASCVD risk assessment tool and other key biomarkers of AD and cognitive deterioration. This will provide a more comprehensive understanding of the complex interplay between these factors in AD initiation.

## Data Availability

Data are available upon reasonable request.

## Notes

### Competing Interest Statement

The authors have declared no competing interest.

### Funding Statement

SOURCES OF FUNDING This work was funded by grants R01-AG072641, R01-AG052446 and R01-AG064877 from the National Institute on Aging. The content is solely the responsibility of the authors and does not necessarily represent the official views of the National Institutes of Health.

### Author Declarations

Institutional Review Board permission by the University of Pittsburgh was obtained. CONSENT STATEMENT All human participants provided informed consen

